# Dual Cannabis-Methamphetamine Use Doubles Bipolar Disorder Risk After Psychosis: A 25-Year Inpatient Cohort Study from Iran

**DOI:** 10.1101/2025.08.27.25334535

**Authors:** Zeinab Khodaparast, Peyman Saeedi, Kobra Hosseini, Haniyeh Rashidi, Mahin Seifi alan, Mahya Dorri, Farzaneh Mirsharifi, Elham Molaei Birgani, Hadith Rastad

**Affiliations:** Psychiatry Department, Kazae43Avosh Cognitive Behaviour Sciences and Addiction Research Center, Shafa educational - remedial Hospital, Guilan University of Medical Sciences, Rasht, Iran; Clinical Research and Development Center of the Kamali Hospital, Alborz University of Medical Sciences, Karaj, Iran; Cardiovascular Research Center, Alborz University of Medical Sciences, Karaj, Iran; Department of Obstetrics & Gynecology and Health Research Methods, Evidence and Impact, McMaster University, Hamilton, Ontario, Canada; Family health division, Tehran northwest 2 primary health network, Iran University of Medical Sciences, Tehran, Iran

**Author notes:** Corresponding authors: Hadith Rastad. Zeinab Khodaparast and Peyman Saeedi equally contributed as first authors.

**Keywords:** Substance-induced psychosis (SIP), Schizophrenia, Bipolar disorder, Methamphetamine, Cannabis

## Abstract

**Objective:** This study aimed to estimate transition rates from substance-induced psychosis (SIP) to schizophrenia or bipolar disorder, evaluate the risk associated with dual use of methamphetamine and cannabis, and identify predictors of chronicity among Iranian men.

**Methods:** A 25-year retrospective cohort study (1999–2024) was conducted using clinical data from Shafa Hospital, a tertiary psychiatric referral center in Guilan Province, Iran. The study included 258 male inpatients aged ≥17 years with SIP attributed to cannabis and/or methamphetamine use. Patients with a history of schizophrenia or bipolar disorder were excluded. Outcomes were validated using DSM-V criteria, and Kaplan-Meier survival curves and Cox proportional hazards models were employed for analysis.

**Results:** Over a median follow-up of 33 months, 37.6% of participants transitioned to schizophrenia (25.2%) or bipolar disorder (12.4%). The median time to conversion was shorter for bipolar disorder (13.4 months) compared to schizophrenia (26.6 months). Dual cannabis-methamphetamine use significantly increased the risk of bipolar disorder (p=0.008). Familial psychiatric history doubled the risk of schizophrenia (HR=4.29, 95% CI: 2.48–7.41) and tripled the risk of bipolar disorder (HR=3.89, 95% CI: 1.87–8.11). Recurrent hospitalizations were associated with increased risks for both schizophrenia (HR=1.34) and bipolar disorder (HR=2.64). The cumulative incidence of schizophrenia rose linearly to 67.6% at 10 years.

**Conclusion:** SIP poses a significant risk for the development of schizophrenia or bipolar disorder, particularly in cases involving dual-substance use, younger age, familial psychiatric history, and frequent hospitalizations. The findings underscore the need for targeted surveillance, early intervention, and long-term, risk-stratified care models in regions with rising methamphetamine use to reduce SIP-related morbidity

## Introduction

Substance use disorders are an expanding global public health problem, and psychoactive substances contribute significantly to morbidity and mortality worldwide (1–3). In Iran, substance use is disproportionately represented by gender with 24.1% of men having used compared to 2.2% of women(4). Among the severe abstinence complications from substance abuse, Substance-Induced Psychosis (SIP)—an acute psychotic disorder brought on by exposure to drugs—is under criticism regarding its potential transition into chronic psychiatric disease (5–7). While SIP disappears with drug discontinuation, the latest evidence presents that most of the patients progress into chronic psychiatric illnesses like schizophrenia and bipolar illness, challenging the long-established contention of SIP being a self-limited benign sickness(5, 8, 9).

Such individuals are often lost to follow-up and are not usually part of studies (6, 10). Recent literature on SIP transitions is limited and geographically skewed, with most studies being conducted in Europe and North America (6, 11). There remain critical gaps, most prominently in dual substance synergism and regional variation in substance profiles, and considerable controversy for transition rate from SIP to schizophrenia or bipolar disorder and risk factors (6, 12, 13). Methamphetamine trafficking has risen in the Middle East (14), and use of cannabis is on the rise globally (2, 15), but the combined neuropsychiatric hazard of concurrent use of multiple substances by mass numbers of drug users (9), remains underinvestigated. Also, while transition into schizophrenia has been well studied, progression into bipolar disorder is not as well researched, although it is clearly clinically significant (9, 11). Understanding of the process and the factors of risk would be crucial to prevention interventions and managements (6, 10, 12, 13). This retrospective cohort study bridges these gaps by investigating the incidence and predictors of SIP progression to schizophrenia or bipolar disorder in an underrepresented group: Iranian men.

Based on 25 years of clinical data from a tertiary psychiatric referral center, we aim to (1) quantify transition rates to schizophrenia and bipolar disorder, (2) evaluate the synergistic risk of concurrent cannabis and methamphetamine use, and (3) identify demographic, family, and clinical risk factors. By focusing on a high-risk population in a region with increasing methamphetamine trafficking, this study provides new information on long-term SIP outcomes, and establishes an evidence base for the creation of targeted early intervention programs in resource-limited environments.

## Methods

### Study Design and Setting

This retrospective cohort study analyzed clinical data of male patients admitted to the Emergency Department (ED) of Shafa Health Center, a tertiary psychiatric referral hospital in Rasht, Iran. Data were retrieved in April 2024 for a period of 25 years (1999–2024). The study contrasted the frequency of occurrence and predictors of conversion from substance-induced psychosis (SIP) to schizophrenia or bipolar disorder, as characterized based on DSM-V criteria (5).

### Participants

Patients who were included were male patients aged ≥17 years who had a history of one or more Emergency Department (ED) admissions for SIP due to use of cannabis and/or methamphetamine, with the following complete medical records: medical histories, psychiatric assessments, and discharge summaries. Those with existing diagnoses of schizophrenia, bipolar disorder, or schizoaffective disorder before the onset of SIP were excluded. 258 patients were found eligible.

### Variables and Measurements

The study collected information in four main domains: demographic factors (age of initial SIP admission, marital status, employment, education), substance use patterns (primary substances: cannabis-only, methamphetamine-only, or dual; polysubstance history with alcohol and opium), clinical factors (family psychiatric disorder history, history of head trauma, personality disorders), and outcomes (conversion to schizophrenia or bipolar disorder confirmed by two psychiatric assessments).

Dates of both subsequent SIP diagnosis and subsequent psychiatric outcome were recorded to track progression times. Admission patterns were tracked, readmissions two or more ED visits for SIP-related symptoms that occurred at least one week apart. Statistical Analysis

All analysis was performed with R statistical software (version 4.4.2).

Descriptive statistics summarized continuous variables as medians with interquartile ranges (IQRs) and categorical variables as frequencies and percentages. Group differences were established using Mann-Whitney U tests for continuous measures and chi-square or Fisher’s exact tests for categorical measures. Patients with missing data were removed to minimize bias. Statistical significance was a two-tailed P value < 0.05. Subgroup analyses were carried out to evaluate predictors of schizophrenia and bipolar disorder separately. Survival analysis used Kaplan-Meier curves to estimate time-to-conversion by substance type and family history, with log-rank tests between subgroups. Median time-to-conversion with IQRs was calculated in the overall cohort and stratified by diagnostic outcome. Univariable Cox proportional hazards models established predictors of conversion, with hazard ratios (HRs) presented with 95% confidence intervals (CIs). Variables with P < 0.200 in univariable analyses were included in age-adjusted bivariable models, and Schoenfeld residuals ensured proportional hazards assumptions. Analytic code is available upon request to facilitate reproducibility of results. Ethical Considerations

The study was approved by the Shafa Educational & Remedial Center Ethics Committee. The Research and Ethics Committee of Alborz University of Medical Sciences (ABZUMS) reviewed and approved the study proposal and waived the requirement for informed consent. All data were anonymized prior to analysis to preserve patient confidentiality.

### Result

This study examined the conversion rates and predictors to schizophrenia or bipolar disorder following SIP after cannabis and/or methamphetamine dependence in 258 inpatient men followed up for a median of 33.3 months (IQR: 17.9-60.9). (Table 1)

**Table 1:**
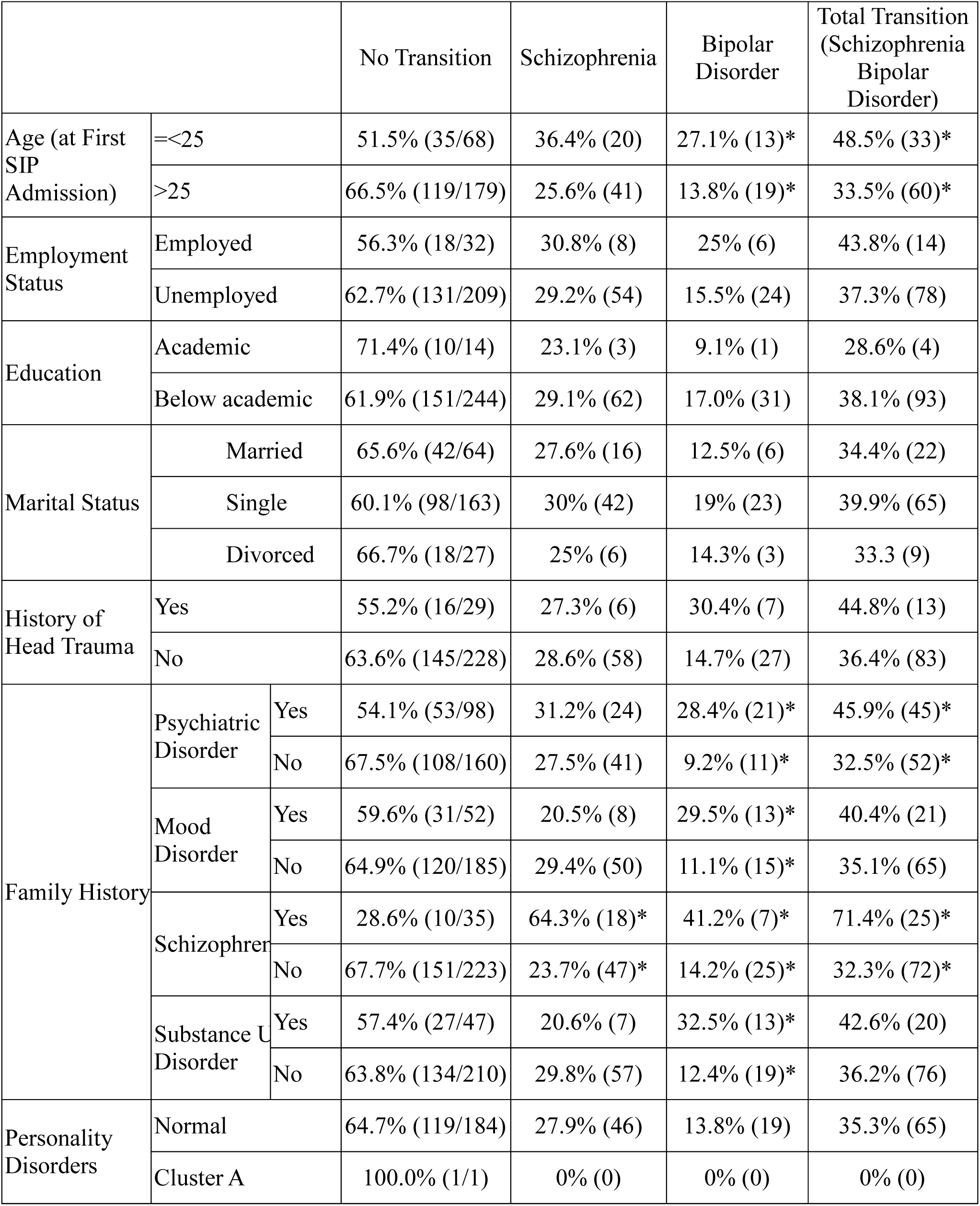
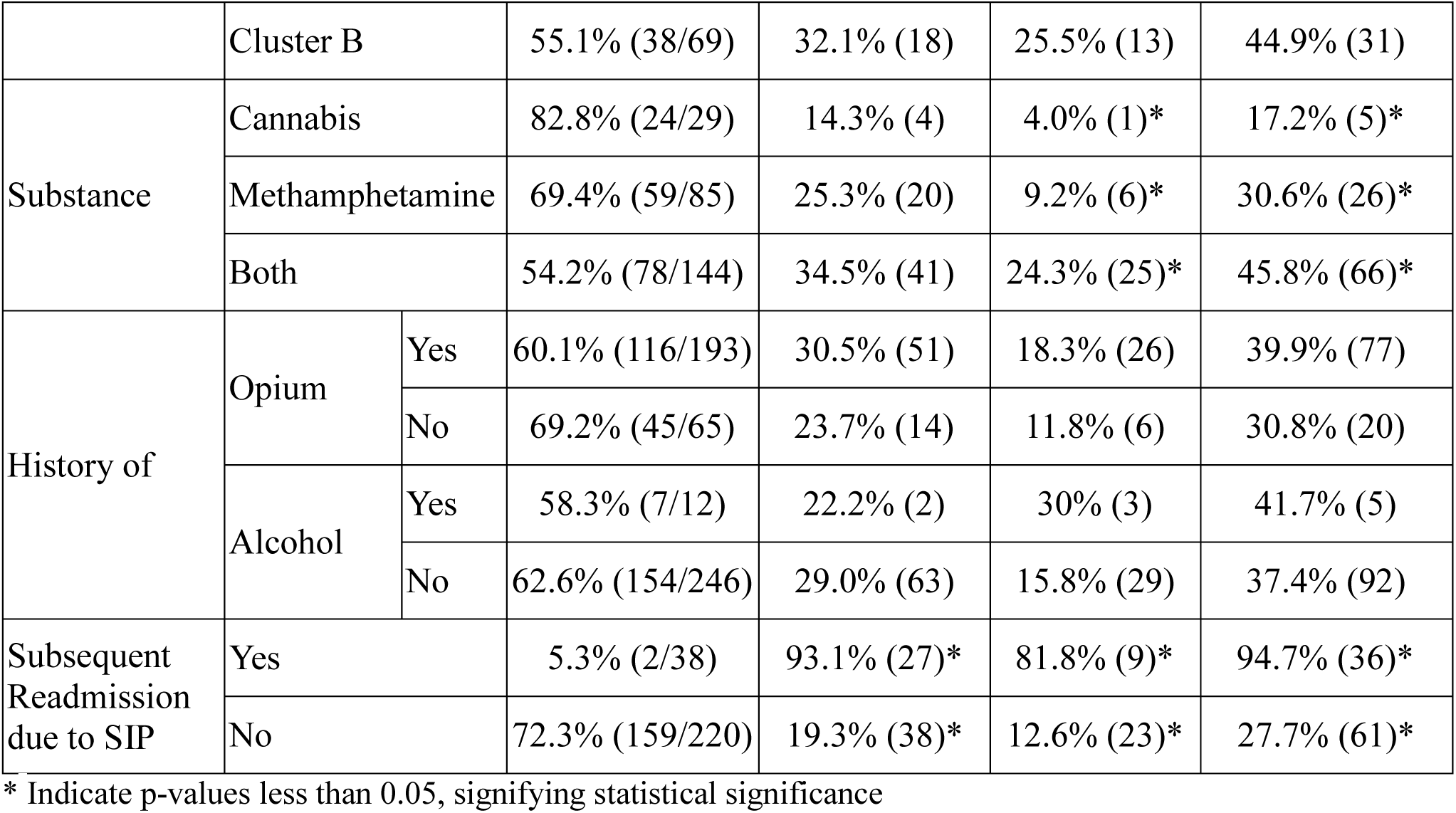
Transition Rates from Substance-Induced Psychosis (SIP) to Schizophrenia or Bipolar Disorder by Baseline Characteristics of Participants.

|  |  |  | No Transition | Schizophrenia | Bipolar Disorder | Total Transition (Schizophrenia Bipolar Disorder) |
| --- | --- | --- | --- | --- | --- | --- |
| Age (at First SIP Admission) | ≤25 |  | 51.5% (35/68) | 36.4% (20) | 27.1% (13)* | 48.5% (33)* |
|  | >25 |  | 66.5% (119/179) | 25.6% (41) | 13.8% (19)* | 33.5% (60)* |
| Employment Status | Employed |  | 56.3% (18/32) | 30.8% (8) | 25% (6) | 43.8% (14) |
|  | Unemployed |  | 62.7% (131/209) | 29.2% (54) | 15.5% (24) | 37.3% (78) |
| Education | Academic |  | 71.4% (10/14) | 23.1% (3) | 9.1% (1) | 28.6% (4) |
|  | Below academic |  | 61.9% (151/244) | 29.1% (62) | 17.0% (31) | 38.1% (93) |
| Marital Status | Married |  | 65.6% (42/64) | 27.6% (16) | 12.5% (6) | 34.4% (22) |
|  | Single |  | 60.1% (98/163) | 30% (42) | 19% (23) | 39.9% (65) |
|  | Divorced |  | 66.7% (18/27) | 25% (6) | 14.3% (3) | 33.3 (9) |
| History of Head Trauma | Yes |  | 55.2% (16/29) | 27.3% (6) | 30.4% (7) | 44.8% (13) |
|  | No |  | 63.6% (145/228) | 28.6% (58) | 14.7% (27) | 36.4% (83) |
| Family History | Psychiatric Disorder | Yes | 54.1% (53/98) | 31.2% (24) | 28.4% (21)* | 45.9% (45)* |
|  |  | No | 67.5% (108/160) | 27.5% (41) | 9.2% (11)* | 32.5% (52)* |
|  | Mood Disorder | Yes | 59.6% (31/52) | 20.5% (8) | 29.5% (13)* | 40.4% (21) |
|  |  | No | 64.9% (120/185) | 29.4% (50) | 11.1% (15)* | 35.1% (65) |
|  | Schizophrenia | Yes | 28.6% (10/35) | 64.3% (18)* | 41.2% (7)* | 71.4% (25)* |
|  |  | No | 67.7% (151/223) | 23.7% (47)* | 14.2% (25)* | 32.3% (72)* |
|  | Substance Use Disorder | Yes | 57.4% (27/47) | 20.6% (7) | 32.5% (13)* | 42.6% (20) |
|  |  | No | 63.8% (134/210) | 29.8% (57) | 12.4% (19)* | 36.2% (76) |
| Personality Disorders | Normal |  | 64.7% (119/184) | 27.9% (46) | 13.8% (19) | 35.3% (65) |
|  | Cluster A |  | 100.0% (1/1) | 0% (0) | 0% (0) | 0% (0) |
|  | Cluster B |  | 55.1% (38/69) | 32.1% (18) | 25.5% (13) | 44.9% (31) |
| Substance | Cannabis |  | 82.8% (24/29) | 14.3% (4) | 4.0% (1)* | 17.2% (5)* |
|  | Methamphetamine |  | 69.4% (59/85) | 25.3% (20) | 9.2% (6)* | 30.6% (26)* |
|  | Both |  | 54.2% (78/144) | 34.5% (41) | 24.3% (25)* | 45.8% (66)* |
| History of | Opium | Yes | 60.1% (116/193) | 30.5% (51) | 18.3% (26) | 39.9% (77) |
|  |  | No | 69.2% (45/65) | 23.7% (14) | 11.8% (6) | 30.8% (20) |
|  | Alcohol | Yes | 58.3% (7/12) | 22.2% (2) | 30% (3) | 41.7% (5) |
|  |  | No | 62.6% (154/246) | 29.0% (63) | 15.8% (29) | 37.4% (92) |
| Subsequent Readmission due to SIP | Yes |  | 5.3% (2/38) | 93.1% (27)* | 81.8% (9)* | 94.7% (36)* |
|  | No |  | 72.3% (159/220) | 19.3% (38)* | 12.6% (23)* | 27.7% (61)* |

\* Indicate p-values less than 0.05, signifying statistical significance

Participants were 31.0 years (IQR: 25.0–36.0) with age ≤25 years in 27.5% (n=68). Educationally, 5.4% (n=14) had academic degrees. Admission diagnoses were dual-substance-induced psychosis (55.8%, n=144), methamphetamine-induced psychosis (32.9%, n=85), and cannabis-induced psychosis (11.2%, n=29). In follow-up, 97 patients (incidence risk: 37.60%, 95% CI:31.67-43.82%) developed schizophrenia or bipolar disorder. Of the transitioned, 65 had schizophrenia (incidence risk: 25.19%, 95% CI:20.02-30.95%), and 32 bipolar disorder (incidence risk: 12.40%, 95% CI:8.64-17.06%). We compared affected participants with their counterparts, no transition group (N=161). The median time from first SIP episode to transition was 20.7 months (IQR: 9.6 - 48.7) that was significantly less in the subjects with family history of psychiatric disorder (13.0 vs. 31.2; P value = 0.019). The median time from first SIP episode to schizophrenia was 26.6 months (IQR: 11.5 – 53.6) (Figure 1), and to bipolar disorder was 13.4 months (IQR: 7.4 – 33.3). (figure.2)

**Fig. 1:**
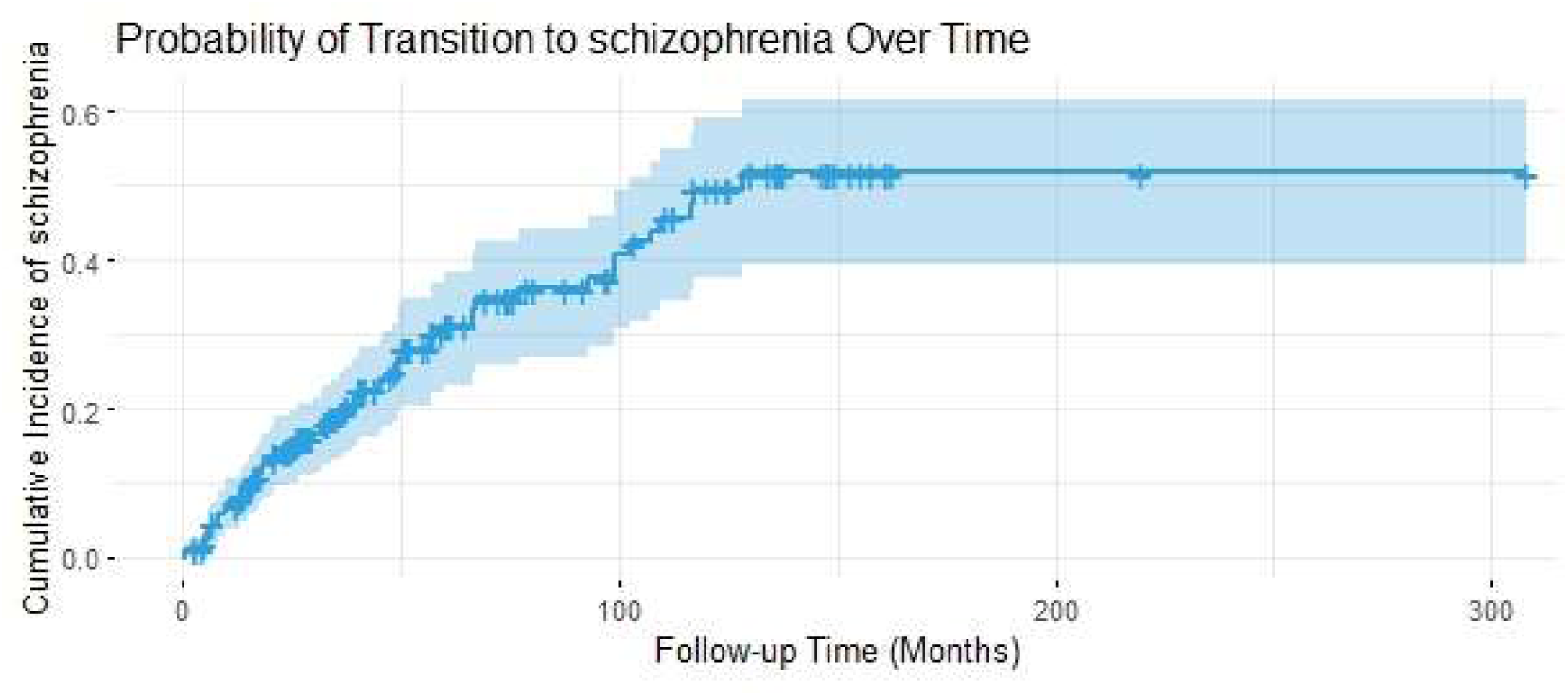
Kaplan-Meier curve illustrating the cumulative probability of transition from substance-induced psychosis (SIP) to schizophrenia over follow-up time (in months). The probability of transition rises steadily, with a median time to conversion of 26.6 months. Familial psychiatric illness and recurrent admissions were significant predictors of schizophrenia transition.

**Fig. 2:**
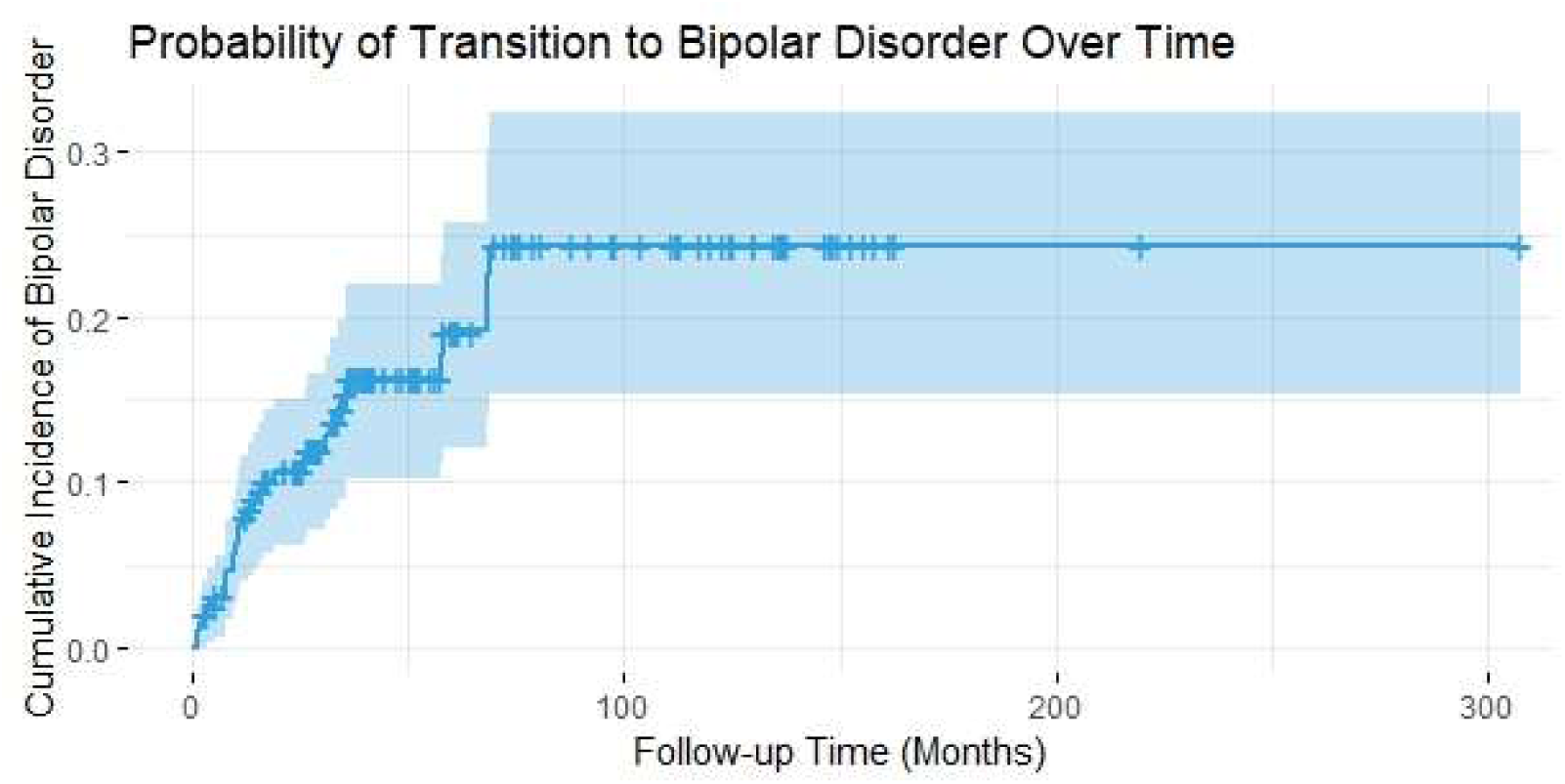
Kaplan-Meier curve showing the cumulative probability of transition from substance-induced psychosis (SIP) to bipolar disorder over follow-up time (in months). The probability of transition increases over time, with a median time to conversion of 13.4 months. Dual cannabis-methamphetamine use significantly increased the risk of bipolar disorder (p=0.008).

The cumulative incidence (Supplementary Table.1) of schizophrenia increased steadily during follow-up from 15.5% at 2 years to 67.6% at 10 years.

On the other hand, cumulative incidence for bipolar disorder plateaued at 27.7% after Year 6. Cumulative incidence of transition rates (bipolar disorder or schizophrenia) rose from 22.9% at 2 years to 86.7% at 10 years, crossing 50% in Year 5 and 90% in Year 14. Furthermore, the cumulative incidence of schizophrenia transitions remained consistently higher than bipolar disorder with a difference in cumulative hazards at Year 10 of 40.1% (67.6% vs. 27.7%). (Figure.3). Cumulative incidence for conversion from SIP to schizophrenia and/or bipolar disorder Younger age of first SIP admission (≤25 years) was associated with higher rates of conversion to bipolar disorder (27.1% vs 13.8% in older patients; P value = 0.046), but for conversion to schizophrenia it did not reach significancy.

**Fig. 3:**
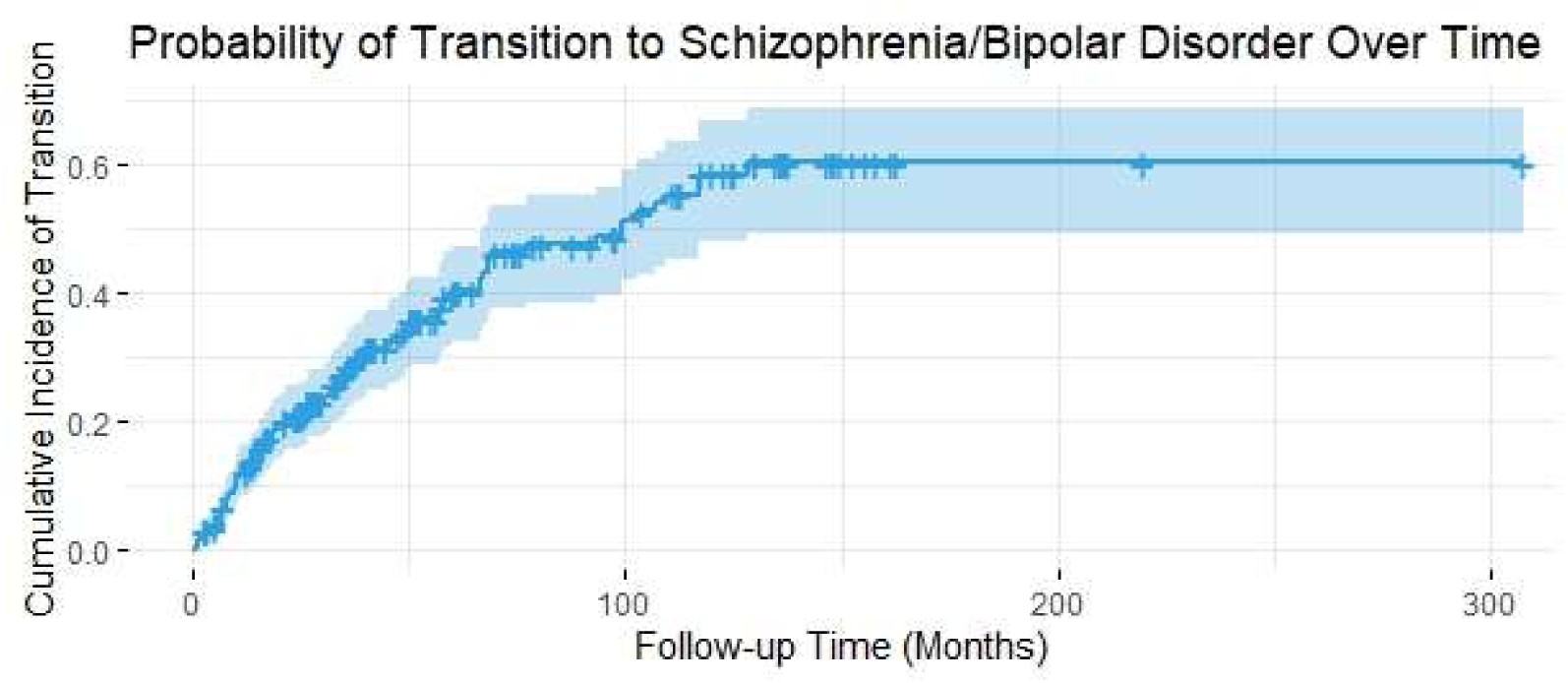
Comparative Kaplan-Meier curves depicting the cumulative probability of transition from substance-induced psychosis (SIP) to schizophrenia and bipolar disorder over follow-up time (in months). The cumulative incidence of schizophrenia transitions (67.6% at 10 years) was consistently higher than that of bipolar disorder transitions (27.7% at 10 years). Dual-substance use and family psychiatric history were key predictors of transition.

Family history of psychiatric disorders raised the risk of transition to bipolar disorder (28.4% vs. 9.2%; P value = 0.001), and family history of mood disorders (29.5% vs. 11.1%; P value = 0.007) and substance abuse disorders (32.5% vs. 12.4% P value = 0.007) were also true.

These three associations were statistically not significant for transition to schizophrenia.

Further, family history of schizophrenia demonstrated correlation with both conditions: schizophrenia (64.3% vs. 23.7%; P value < 0.001), and bipolar disorder (41.2% vs. 14.2%; P value = 0.010). A considerably higher transition rate to bipolar disorder was observed in patients who consumed both methamphetamine and cannabis (24.3%) compared to only-methamphetamine (9.2%) and only-cannabis (4.0%) consumers with a P value of 0.008. The association was not significant for transition to schizophrenia but not to total transition (P value = 0.008). Subsequent readmissions for schizophrenia SIP had higher transition to schizophrenia (93.1% vs. 19.3%; P value < 0.001), and for bipolar disorder (81.8% vs. 12.6%; P value < 0.001). No relations were significant regarding employment status, education level, marital status, history of head trauma, and histories of abuse of opium or alcohol, or personality disorders (P value > 0.05).

### Unadjusted Hazard Ratios for Transition

In univariable Cox regression analysis, the subjects with early age at first SIP admission (≤25 years) were 2.13 times more likely to develop into bipolar disorder (HR = 2.13, 95% CI: 1.05– 4.31).

Increased risk for bipolar disorder was observed with familial psychiatric disorder threefold (HR = 3.89, 95% CI: 1.87–8.11), particularly family history of mood disorder (HR = 3.21, 95% CI: 1.52–6.78), and substance use disorder (HR = 3.67, 95% CI: 1.79–7.50). (Table 2)

**Table 2:**
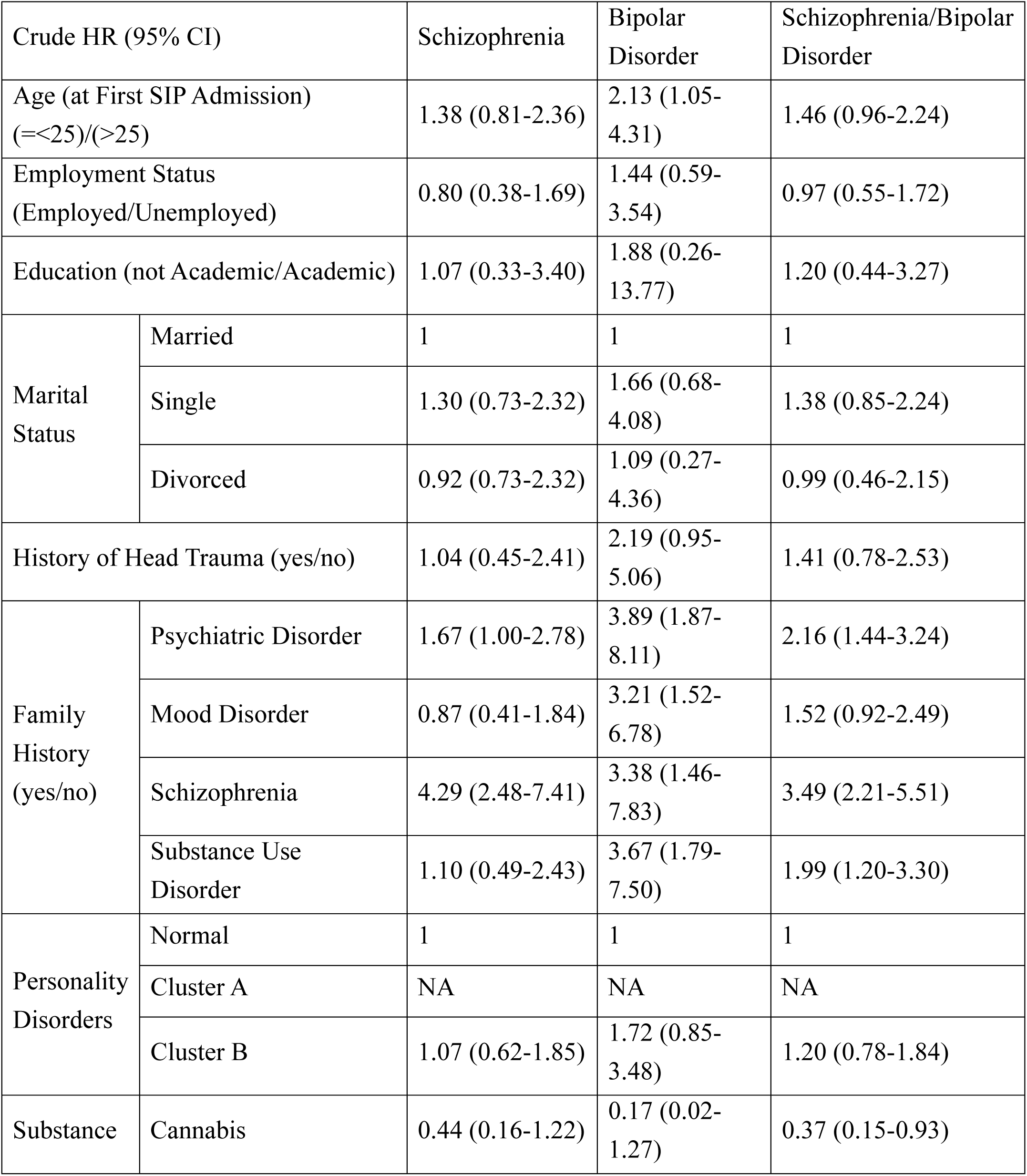
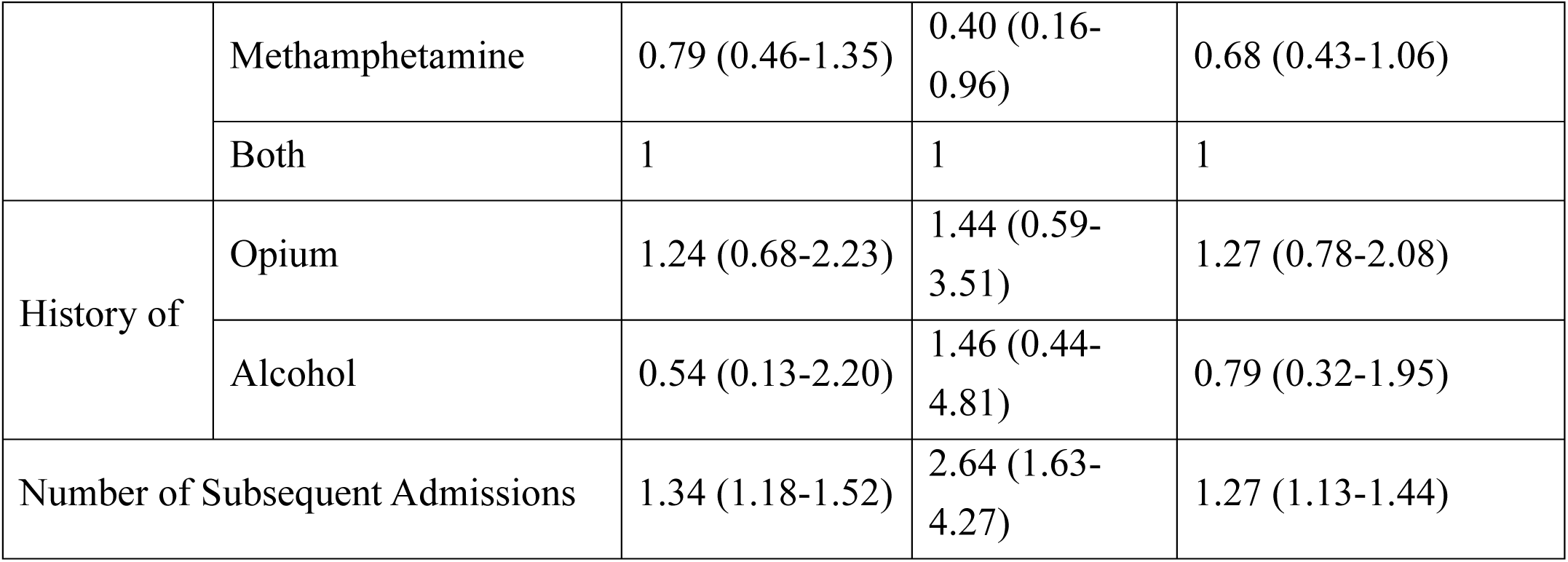
Univariate Cox Regression Analysis of Hazard Ratios for Transition from Substance-Induced Psychosis (SIP) to Schizophrenia or Bipolar Disorder.

| Crude HR (95% CI) |  | Schizophrenia | Bipolar Disorder | Schizophrenia/Bipolar Disorder |
| --- | --- | --- | --- | --- |
| Age (at First SIP Admission) (=≤25)/(>25) |  | 1.38 (0.81-2.36) | 2.13 (1.05-4.31) | 1.46 (0.96-2.24) |
| Employment Status (Employed/Unemployed) |  | 0.80 (0.38-1.69) | 1.44 (0.59-3.54) | 0.97 (0.55-1.72) |
| Education (not Academic/Academic) |  | 1.07 (0.33-3.40) | 1.88 (0.26-13.77) | 1.20 (0.44-3.27) |
| Marital Status | Married | 1 | 1 | 1 |
|  | Single | 1.30 (0.73-2.32) | 1.66 (0.68-4.08) | 1.38 (0.85-2.24) |
|  | Divorced | 0.92 (0.73-2.32) | 1.09 (0.27-4.36) | 0.99 (0.46-2.15) |
| History of Head Trauma (yes/no) |  | 1.04 (0.45-2.41) | 2.19 (0.95-5.06) | 1.41 (0.78-2.53) |
| Family History (yes/no) | Psychiatric Disorder | 1.67 (1.00-2.78) | 3.89 (1.87-8.11) | 2.16 (1.44-3.24) |
|  | Mood Disorder | 0.87 (0.41-1.84) | 3.21 (1.52-6.78) | 1.52 (0.92-2.49) |
|  | Schizophrenia | 4.29 (2.48-7.41) | 3.38 (1.46-7.83) | 3.49 (2.21-5.51) |
|  | Substance Use Disorder | 1.10 (0.49-2.43) | 3.67 (1.79-7.50) | 1.99 (1.20-3.30) |
| Personality Disorders | Normal | 1 | 1 | 1 |
|  | Cluster A | NA | NA | NA |
|  | Cluster B | 1.07 (0.62-1.85) | 1.72 (0.85-3.48) | 1.20 (0.78-1.84) |
| Substance | Cannabis | 0.44 (0.16-1.22) | 0.17 (0.02-1.27) | 0.37 (0.15-0.93) |
|  | Methamphetamine | 0.79 (0.46-1.35) | 0.40 (0.16-0.96) | 0.68 (0.43-1.06) |
|  | Both | 1 | 1 | 1 |
| History of | Opium | 1.24 (0.68-2.23) | 1.44 (0.59-3.51) | 1.27 (0.78-2.08) |
|  | Alcohol | 0.54 (0.13-2.20) | 1.46 (0.44-4.81) | 0.79 (0.32-1.95) |
| Number of Subsequent Admissions |  | 1.34 (1.18-1.52) | 2.64 (1.63-4.27) | 1.27 (1.13-1.44) |

While dual use of drugs (methamphetamine and marijuana) served as the reference category, use of only methamphetamine was associated with reduced risk of bipolar disorder compared to the reference category (HR = 0.40, 95% CI: 0.16–0.96).

Each additional readmission for SIP raised the risk of bipolar disorder by 164% (HR = 2.64, 95% CI: 1.63–4.27). Family psychiatric history slightly increased the risk for schizophrenia (HR = 1.67, 95% CI: 1.00–2.78), but schizophrenia family history doubled the hazard (HR = 4.29, 95% CI: 2.48–7.41), although less than 25 years was not a predictor for schizophrenia. Each SIP readmission increasingly elevated the risk for schizophrenia by 34% (HR = 1.34, 95% CI: 1.18– 1.52). Cannabis use alone lowered overall transition risk compared with dual use (HR = 0.37, 95% CI: 0.15–0.93) All statistically significant correlations were still substantial in age-adjusted bivariable Cox regression analyses, except two associations: (1) between psychiatric disorder family history and conversion to schizophrenia, and (2) between methamphetamine-only use and conversion to bipolar disorder.

Hazard ratios for the transition from SIP to psychotic disorder in the univariate COX regression model

## Discussion

This 258 Iranian male inpatient retrospective cohort study of substance-induced psychosis (SIP) due to cannabis and/or methamphetamine identified a 37.6% transition rate to schizophrenia (25.2%) or bipolar disorder (12.4%) over a median follow-up of 33 months.

Key predictors included family history of schizophrenia and multiple SIP readmissions for schizophrenia; early age at first SIP admission (≤25 years), family histories of psychiatric-, mood-, and substance use disorders, dual cannabis-methamphetamine use, and multiple SIP readmissions for bipolar disorder.

These findings are in contrast to the perception of SIP as a transient condition and suggest its value as a marker of high risk of chronic mental illness.

The rate of transition of schizophrenia (25.2%) is in line with meta-analytic returns (22–28%) but greater than lower rates reported in some European research (6–17%) (6, 9, 11, 16–18). These discrepancies could be explained by regional differences in drug profiles, follow-up times, and study designs. Our inpatient sample, which consists of severe cases, also could overestimate transition rates compared with population registries.

For bipolar disorder, the 12.4% transition rate exceeds prior estimates (4.5–8.4%) (9, 11), potentially due to our focus on dual cannabis-methamphetamine use, which elevated bipolar disorder risk to 24.3%. Cumulative incidence of schizophrenia rose steadily from 15.5% at 2 years to 67.6% at 10 years, but the levelled off at 27.7% for bipolar disorder after Year 6. This reflects distinct disease processes and suggests that follow-up for at least many years, particularly among those at highest risk such as dual-substance users or psychiatrically ill parents, is paramount. The increasing trend of chronic incidence of schizophrenia aligns with studies of Norwegian (9) and Danish cohorts (11). Methamphetamine and cannabis were similarly high-risk drugs, with comorbid consumption increasing susceptibility to bipolar disorder. This concurred with existing research linking cannabis-induced psychosis with a higher incidence of schizophrenia transition (34–47.4%) and amphetamines with an elevated risk of psychosis (22–30%) (6, 9, 11, 17, 18, 20). What this study reveals is the need for classification of substances at a fine-grained level in order to assess risk profiles, as many studies aggregate different drug combinations and reduce specificity (6, 9, 18, 20). Younger age (≤25 years) predicted bipolar disorder transitions (HR = 2.13), contrary to studies linking youth to risk for schizophrenia (9, 11, 16–18, 20). Adolescence and early adulthood are critical stages of brain development, and drug exposure during this stage may disrupt neurodevelopmental processes, particularly exposing younger individuals to risk for bipolar disorder pathways (22–26).

Family history of schizophrenia increased the risk of schizophrenia fourfold (HR = 4.29), while family history of mood and substance use disorders predicted bipolar disorder outcomes independently. These findings align with genetic liability models and reinforce the value of family history as a crucial screening tool (17). Kendler et al. (2019) demonstrated that familial psychosis liability predicts conversion, hence making family history a significant screening tool (17).

Repeated SIP readmissions also predicted schizophrenia (HR = 1.34) and bipolar disorder (HR = 2.64), highlighting shortcomings in post-discharge care. The dose-response relationship between readmission frequency and diagnostic progression underlies systemic interventions to prevent recurrent psychosis and emergency department utilization (11, 27–29).

Median transition times were shorter for bipolar disorder (13.4 months) than for schizophrenia (26.6 months), in contrast to some European research (9, 11). This suggests population-specific progression patterns, perhaps based on substance profiles or healthcare access.

SIP may take place in drug-abuse-prone patients with elevated familial risk for drug abuse and intermediate risk for psychosis. Repeated SIP admissions may signify substance-induced recurrences or incipient, unsuspected phases of schizophrenia, challenging the classical discrimination between substance-induced and primary psychiatric disorders (6, 12, 17, 30).

Strengths include well-defined cohort, high-resolution substance classification, and long follow-up. Limitations include single-center recruitment, cohort of males only, use of clinical charts, and irregular follow-up periods. Absence of frequency and dosage data rules out dose-response analyses (6, 10).

Clinically, the implications of these findings highlight risk stratification, early extended follow-up, drug-specific treatments, enhanced post-discharge care, and family-oriented psychoeducation. The inclusion of SIP patients within early multimodal psychosis treatment pathways and maximizing personalized treatments can prevent long-term morbidity (6, 10, 17, 19, 29).

In summary, this research calls attention to the high risk of chronic mental disease after SIP, with 37.6% developing schizophrenia or bipolar disorder. Dual-substance use, family psychiatric history, and multiple admissions were the critical predictors. The findings contradict the view of SIP as a fleeting phenomenon and underline the importance of early, holistic treatment to forestall long-term morbidity

## Data Availability

All data produced in the present study are available upon reasonable request to the authors

## Acknowledgements

The authors acknowledge the Clinical Research Development Center of Kamali and Rajaee Hospitals (Alborz University of Medical Sciences) for administrative and technical support.

## Authors’ information

Not applicable.

## Clinical trial number

Not applicable. This is not a clinical trial.

---

**Supplementary Table 1:** Cumulative Incidence of Transition from Substance-Induced Psychosis (SIP) to Schizophrenia and/or Bipolar Disorder Over Time.

|  | **Cumulative Incidence** |  |  |
| --- | --- | --- | --- |
| **Duration** | **Total Transition** | **Schizophrenia** | **Bipolar Disorder** |
| 2 years | 22.9% | 15.5% | 11.3% |
| 3 years | 33.5% | 21.0% | 17.7% |
| 4 years | 40.6% | 28.9% | 17.7% |
| 5 years | 51.2% | 37.1% | 21.2% |
| 6 years | 61.6% | 42.5% | 27.7% |
| 7 years | 63.7% | 44.6% | 27.7% |
| 8 years | 66.0% | 46.9% | 27.7% |
| 10 years | 86.7% | 67.6% | 27.7% |
| 14 years | 91.2% | 72.1% | 27.7% |

